# Effects of Confounding Bias in COVID-19 and Influenza Vaccine Effectiveness Test-Negative Designs Due to Correlated Influenza and COVID-19 Vaccination Behaviors

**DOI:** 10.1101/2021.10.22.21265390

**Authors:** Margaret K. Doll, Stacy M. Pettigrew, Julia Ma, Aman Verma

## Abstract

**Background:** The test-negative design is commonly used to estimate influenza and COVID-19 vaccine effectiveness (VE). In these studies, correlated COVID-19 and influenza vaccine behaviors may introduce a confounding bias where controls are included with the other vaccine-preventable acute respiratory illness (ARI). We quantified the impact of this bias on VE estimates in studies where this bias is not addressed.

**Methods:** We simulated study populations under varying vaccination probabilities, COVID-19 VE, influenza VE, and proportions of controls included with the other vaccine-preventable ARI. Mean bias was calculated as the difference between true and estimated VE. Absolute mean bias in VE estimates was classified as low (<10%), moderate (10% to <20%), and high (≥20%).

**Results:** Where vaccination probabilities are positively correlated, COVID-19 and influenza VE test-negative studies with influenza and SARS-CoV-2 ARI controls, respectively, underestimate VE. For COVID-19 VE studies, mean bias was low for all scenarios where influenza represented ≤50% of controls. For influenza VE studies, mean bias was low for all scenarios where SARS-CoV-2 represented ≤10% of controls. Although bias was driven by the conditional probability of vaccination, low VE of the vaccine of interest and high VE of the confounding vaccine increase its magnitude.

**Conclusions:** Where a low percentage of controls are included with the other vaccine-preventable ARI, bias in COVID-19 and influenza VE estimates is low. However, influenza VE estimates are likely more susceptible to bias. Researchers should consider potential bias and its implications in their respective study settings to make informed methodological decisions in test-negative VE studies.

## Introduction

Phase four observational studies are essential to examine the direct effects of vaccination in a real-world setting. Due to its relative simplicity, the test-negative study design is the predominant observational design used to estimate vaccine effectiveness (VE) for influenza,^1^ and increasingly, COVID-19.^2-4^ In these studies, test-negative participants are persons who seek healthcare for an acute respiratory illness (ARI) and are tested for the disease of interest. Participants who test positive are classified as “cases”, while participants who test negative are classified as “controls”. VE is estimated using the formula (1 − *odds ratio*) *\** 100, where the odds ratio compares the vaccination odds between cases and controls.^5^

Similar to other case-control studies, controls in the test-negative design are used as a proxy to estimate the true vaccination odds in the source population of cases.^5, 6^ Since COVID-19 and influenza VE test-negative designs select ARI controls who are negative for the disease of interest, a foundational assumption of these designs is that the risks of alternative causes of ARI are independent of exposure status (*i*.*e*., vaccination).^7^ Where this assumption is violated, VE estimates will be biased unless independence is established by deconfounding either in the statistical analysis or the modification of the study design.

To date, work examining the validity of this assumption in influenza test-negative VE studies has focused on direct, biological mechanisms by which influenza and/or influenza vaccination may influence the risk of alternate causes of ARI.^8, 9^ However, a relationship between influenza vaccination and alternate ARI need not be causally-related to violate this assumption; in fact, a violation can also occur due to a relationship established by an indirect, confounding pathway.^10^

Recent systematic reviews and surveys have demonstrated a positive correlation between influenza and COVID-19 vaccination probabilities.^11-14^ Because of this relationship, the risks of influenza and SARS-CoV-2 are no longer independent of the vaccination probabilities for the other vaccine-preventable ARI (*i*.*e*., COVID-19 and influenza, respectively). Therefore, where test-negative controls in either influenza or COVID-19 VE studies include persons with the other vaccine-preventable ARI, vaccination for these diseases acts as a confounder. In these studies, where this confounder is unaccounted for, the fundamental assumption of exposure independence in control selection is violated, leading to bias in VE estimates.

Since SARS-CoV-2 and influenza are likely to co-circulate in upcoming influenza seasons, it is important to understand the scope and magnitude of this confounding bias on COVID-19 and influenza VE estimates. Here, we aim to contribute to this knowledge by: (i) examining its theoretical basis and deconfounding methods to remove bias, and (ii) quantifying bias in COVID-19 and influenza VE estimates using simulations, where this bias is not otherwise addressed.

## Methods

### Theoretical basis

In Figure 1a, we examine the theoretical basis for bias from inclusion of controls with a non-independent exposure in COVID-19 or influenza VE test-negative designs. In the directed acyclic graph (DAG), COVID-19 vaccination *V*_*covid*_ and influenza vaccination *V*_*flu*_ are both related to a common ancestor, which we refer to as an individual’s motivation to seek vaccination (*M*). Here, we use (*M*) to represent a set of unobserved variables, including: (i) beliefs and acceptance of vaccines; (ii) external vaccination pressures that influence the uptake of both vaccines, such as vaccine mandates or policies; and (iii) perceived vulnerability/risks of vaccine-preventable diseases to one-self or vulnerable contacts. Through this common ancestor, a correlated relationship is established between influenza and COVID-19 vaccination probabilities through the pathway *V*_*flu*_ ← (*M*)→ *V*_*covid*_; since each vaccine is also directly related to infection with medically-attended ARI for that disease, the relationships extend to their causal descendants creating the confounding pathways: (i) *V*_*flu*_ ← (*M*)→ *I*_*SARS*−*CoV*−2_, which violates the underlying assumption of exposure independence in influenza VE test-negative designs that include SARS-CoV-2 controls, and (ii) *V*_*covid*_ ← (*M*)→ *I*_*flu*_, which violates this assumption in COVID-19 VE test-negative designs that include influenza controls.

**Figure 1.**
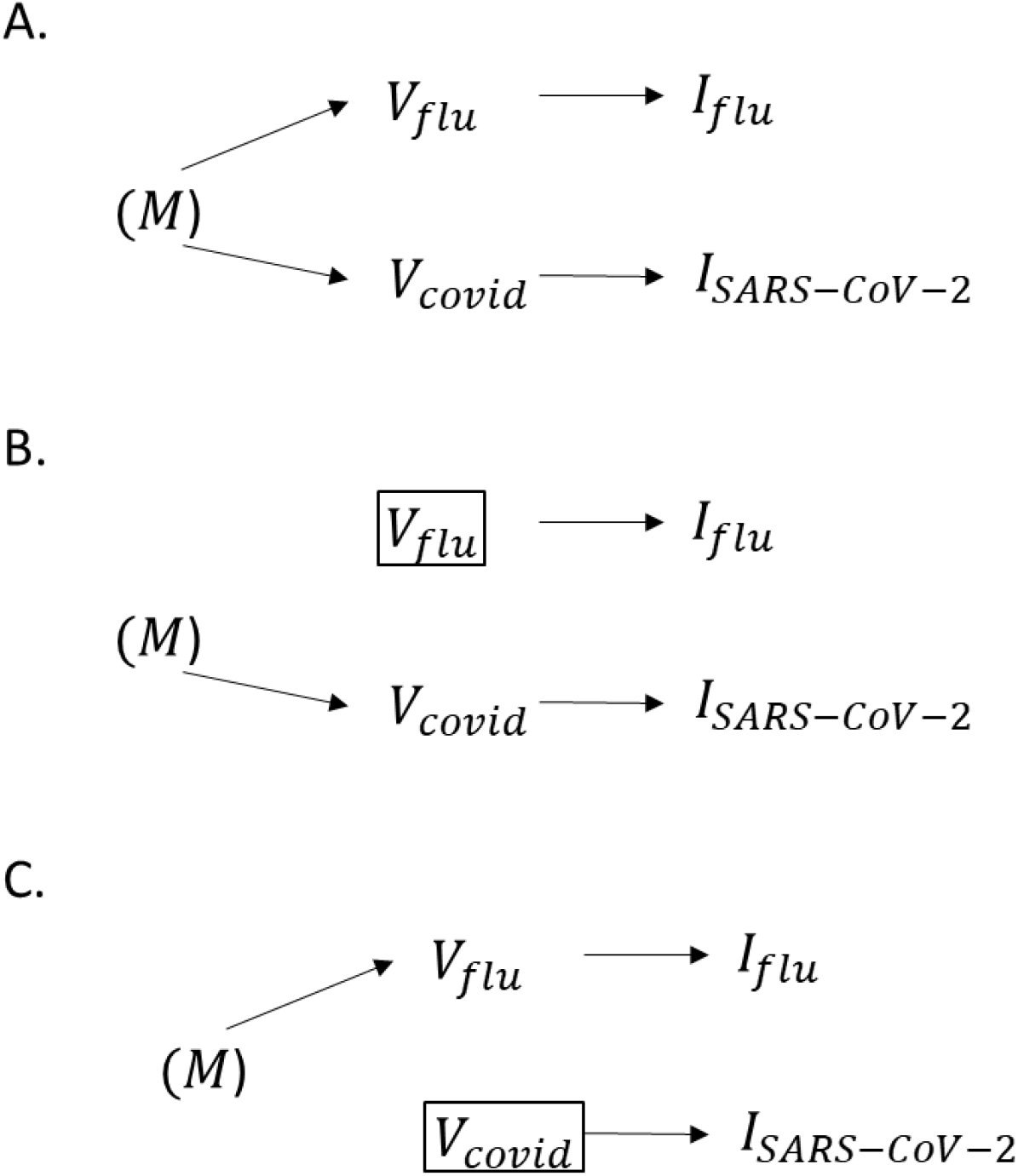
(A) Simplified directed acyclic graph (DAG) illustrating the relationship between COVID-19 and influenza vaccination probabilities. Vaccination motivation *M* is a common ancestor of influenza vaccine uptake *V*_*flu*_ and COVID-19 vaccine uptake *V*_*covid*_. The parentheses indicate that *M* is unmeasured. Through *M* a forked, confounding pathway exists linking *V*_*covid*_ to medically-attended influenza ARI *I*_*flu*_ (*V*_*covid*_ ← (*M*)→ *I*_*flu*_), and *V*_*flu*_ to medically-attended SARS-CoV-2 ARI *I*_*SARS*−*CoV*−2_ (*V*_*flu*_ ← (*M*)→ *I*_*SARS*−*CoV*−2_). (B) Adjustment in the statistical model for *V*_*flu*_ closes the confounding pathway from *V*_*covid*_ ← (*M*)→ *I*_*flu*_ in COVID-19 vaccine effectiveness (VE) test-negative studies that include influenza controls. (C) Similarly, adjustment for *V*_*covid*_ in an influenza VE test-negative study that includes SARS-CoV-2 controls closes the confounding pathway from *V*_*flu*_ ← (*M*)→ *I*_*SARS*−*CoV*−2_.

### Mitigation of Bias in VE estimates

Given the confounding structure in Figure 1a, we propose two potential options to mitigate confounding bias in COVID-19 and influenza test-negative designs, where co-circulation of SARS-CoV-2 and influenza is present: (i) deconfounding in the analysis, or (ii) deconfounding in the study design. Alternatively, a third potential option is to ignore the bias if it is anticipated to be sufficiently small, and not meaningful to VE estimates. We address options (i) and (ii) in the following section, and in subsequent sections, quantify bias using simulations to understand the implications of option (iii).

#### (i) Deconfounding in the analysis

In Figure 1b, we demonstrate how the assumption of exposure independence can be restored in a COVID-19 VE study analysis by statistical adjustment or stratification for influenza vaccination. Similarly, in Figure 1c, we demonstrate how statistical adjustment for COVID-19 vaccination in the analysis can also restore the validity of this assumption in influenza VE studies. Although these mechanisms can recover unbiased estimates of VE, it is important to note that they may interfere with the “efficiency principle” in case-control designs proposed by Wacholder et al.^15^ Where *V*_*covid*_ and *V*_*flu*_ are highly correlated, statistical adjustment for the confounder *V*_*covid*_ in an influenza VE study, or *V*_*flu*_ in a COVID-19 VE study, also reduces the conditional variability of the exposure in the strata of the confounder.^15^ Therefore, although adjustment removes bias, precision of VE estimates may also be reduced.^15^ This may be of particular concern in studies that aim to explore VE among subpopulations of interest or the effect of waning VE, which require additional statistical power.

#### (ii) Deconfounding in the design

As an alternative to deconfounding in the analysis, deconfounding may be achieved in the design through study restriction. To avoid violation of Wacholder et al.’s^15^ efficiency principle here, ample controls must exist that can be enrolled who are independent of the exposure probability. In the setting of a test-negative design, restriction may be implemented by excluding: (i) influenza controls from COVID-19 test-negative VE designs, or (ii) SARS-CoV-2 controls from influenza test-negative VE designs. Practically, these can be achieved by testing participants for both diseases, and only enrolling persons who test negative for <u>both</u> diseases as controls. Another alternative method of restriction is to enroll controls who test positive for a different cause of ARI, which is presumed independent of the exposure probability.

### Simulations to quantify bias in COVID-19 and influenza VE estimates

As previously mentioned, a third option is to ignore bias if it is anticipated to be sufficiently small and not meaningful to VE estimates. However, it is important to understand the potential magnitude of confounding bias to make this determination. We used simulated populations where COVID-19 and influenza vaccination were positively correlated to estimate mean bias in COVID-19 and influenza VE estimates. While simulations were performed for each disease separately, the following common input parameters were included in both analyses:

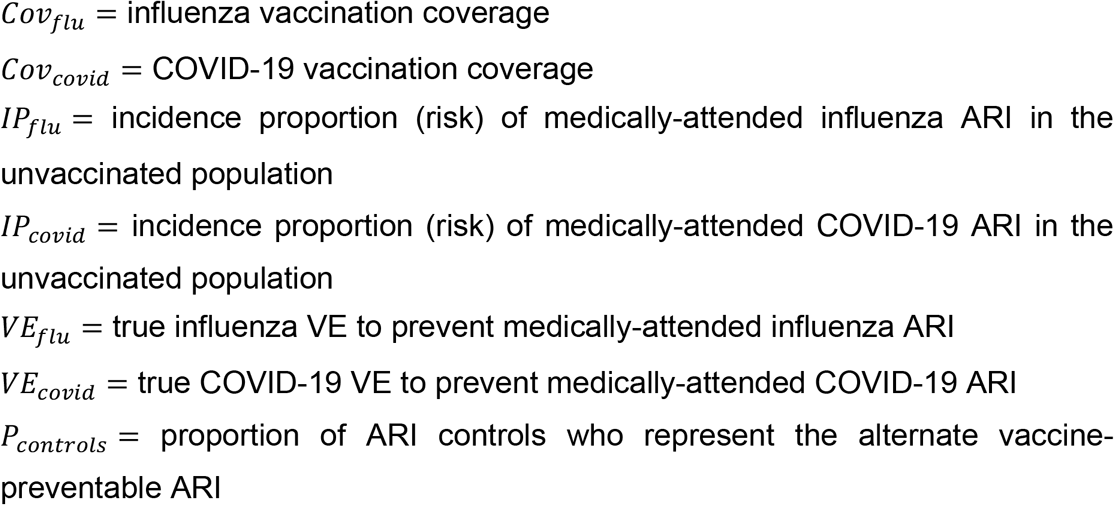

For all simulations, we assumed *Cov*_*flu*_= 55% and *Cov*_*covid*_ = 70%, which approximates 2020-21 influenza vaccination coverage and fully-vaccinated COVID-19 vaccine coverage among U.S. adults.^16, 17^ We also assumed *IP*_*flu*_ = 5% and *IP*_*covid*_ = 5%, based upon previous simulations investigating bias in test-negative designs.^1^ We examined three scenarios for *VE*_*flu*_ of 40%, 50%, and 60% effectiveness against medically-attended influenza ARI, consistent with *VE*_*flu*_ data from recent influenza seasons.^18^ Furthermore, for *VE*_*covid*_, we examined the 3 scenarios of 70%, 80%, and 90% effectiveness against medically-attended SARS-CoV-2 ARI, similar to estimates from recent VE studies.^4, 19-22^ For each scenario, we examined a range for *P*_*controls*_ of 0, 0.1, 0.25, 0.5, 0.75, 0.9 and 1.0.

In addition to these inputs, we included an input variable representing the conditional probability of vaccination given vaccination with the other ARI vaccine. Specifically, *RR*_*Covid vx*|*flu vx*_ was used to simulate bias in COVID-19 VE studies, and *RR*_*flu vx*|*covid vx*_ was used to simulate bias in influenza VE studies. These variables were defined as the following:

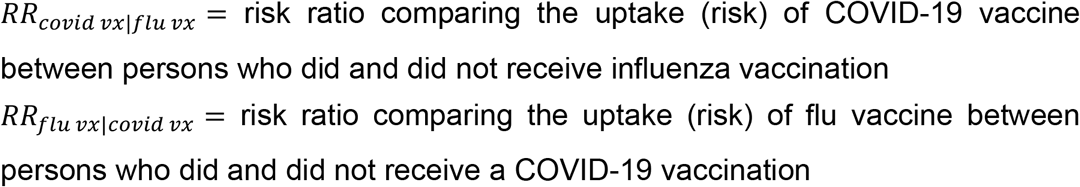

We used a range of input values for *RR*_*covid vx*|*flu vx*_ of 1.5, 2.0, and 3.0; a range of 2.0, 5.0, and 8.0 was used for *RR*_*flu vx*|*covid vx*_. With the exception of *RR*_*covid vx*|*flu vx*_ = 3.0, these estimates were based on the conditional probabilities of influenza and COVID-19 vaccination from a recent, nationally-representative survey of adults in the United States (U.S.) sponsored by the Centers for Disease Control and Prevention (CDC) (Supplemental Appendix 1).^14^ A value of *RR*_*covid vx*|*flu vx*_ = 3.0 was selected to supplement survey data because it represented the upper limit for this value based on out input values of *Cov*_*flu*_= 55% and *Cov*_*covid*_ = 70%. Since we assumed *Cov*_*flu*_ was lower than *Cov*_*covid*_, a reasonable upper limit of *RR*_*flu vx*|*covid vx*_ could not be estimated.

Simulated populations of COVID-19 and influenza test-negative studies were created using three sequential steps that differed slightly for each disease. Specifically, to explore bias in COVID-19 VE test-negative designs, we simulated: (i) the marginal probabilities of COVID-19 and influenza vaccine uptake in the source population, given *Cov*_*flu*_, *Cov*_*covid*_, and *RR*_*covid vx*|*flu vx*_, (ii) the odds of COVID-19 vaccination among SARS-CoV-2 cases, given *IP*_*covid*_, *VE*_*covid*_, and *Cov*_*covid vx*_, and (iii) the odds of COVID-19 vaccination among influenza controls, given the marginal probabilities from step (i), *IP*_*flu*_, *VE*_*flu*_ and *P*_*controls*_. To explore bias in influenza VE test-negative designs, we modified the three steps to the following: simulation of (i) the marginal probabilities of COVID-19 and influenza vaccination in the source population, given *Cov*_*flu*_, *Cov*_*covid*_, and *RR*_*flu vx*|*covid vx*_, (ii) the odds of influenza vaccination among influenza cases, given *IP*_*flu*_, *VE*_*flu*_, and *Cov*_*flu vx*_, and (iii) the odds of influenza vaccination among SARS-CoV-2 controls, given the marginal probabilities simulated in step (i), *IP*_*covid*_, *VE*_*covid*_, and *P*_*controls*_.

For each scenario, we performed 10,000 simulations with a population of 200,000 subjects. Based on the inputs for *IP*_*Covid*_, this starting population approximated the number of cases in recent COVID-19 test-negative VE studies.^4, 20^ From the simulated populations we estimated 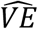 as (1-OR), where OR represents the odds ratio comparing the vaccination odds among cases (simulation step ii) and controls (simulation step iii). Mean bias in VE test negative studies was estimated as the difference of 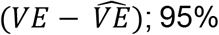 confidence intervals (CI) were calculated as the 2.5^th^ and 97.5^th^ quantiles of the simulated data. We used *a priori* thresholds to classify absolute mean bias of <10% as low, 10% to <20% as moderate, and ≥20% as high. For all parameters where a range of plausible input values were identified, separate populations were simulated for each scenario.

To examine bias associated with only the inclusion of controls with the other vaccine-preventable ARI, we ignored other sources of bias arising from misclassification, unmeasured confounding, and selection bias. All analyses were conducted using RStudio with R version 4.1.0 (The R Foundation for Statistical Computing, Vienna, Austria).

## Results

### Bias in COVID-19 VE estimates

In all scenarios, the inclusion of influenza test-negative controls underestimated true COVID-19 VE. Figure 2 examines mean bias in COVID-19 VE estimates under varying levels of *RR*_*covid vx*|*flu vx*_, *VE*_*flu*_, *VE*_*covid*_, and *P*_*controls*_. In general, there was greater bias in VE estimates with lower *VE*_*covid*_, higher *VE*_*flu*_, increasing *RR*_*covid vx*|*flu vx*_, and increasing influenza *P*_*controls*_. However, in all scenarios, where *P*_*controls*_ with influenza represented ≤50% of the control populations, bias in VE estimates was low (<10%). Bias was also low for all scenarios where *RR*_*covid vx*|*flu vx*_ = 1.5. Moderate to high bias in COVID-19 VE estimates was observed in some scenarios where influenza *P*_*controls*_ approached 75% or more and a *RR*_*covid vx*|*flu vx*_ ≥ 2.0.

**Figure 2.**
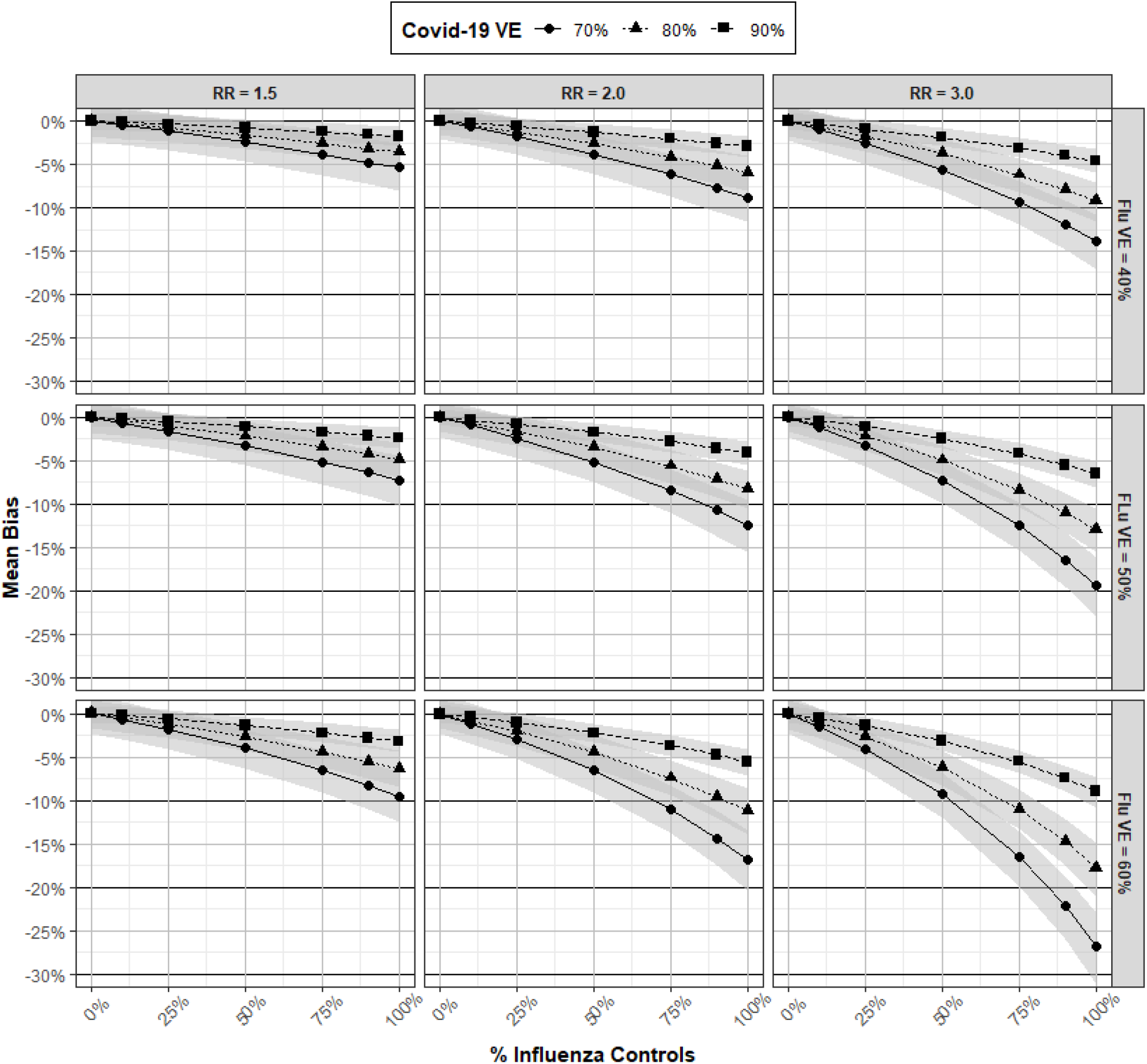
Mean bias and 95% confidence intervals (CI) in COVID-19 vaccine effectiveness (VE) estimates derived from a test-negative study with influenza controls under varying scenarios of *RR*_*covid vx*|*flu vx*_, *VE*_*flu*_, *VE*_*covid*_, and *P*_*controls*_.

**Figure 3.**
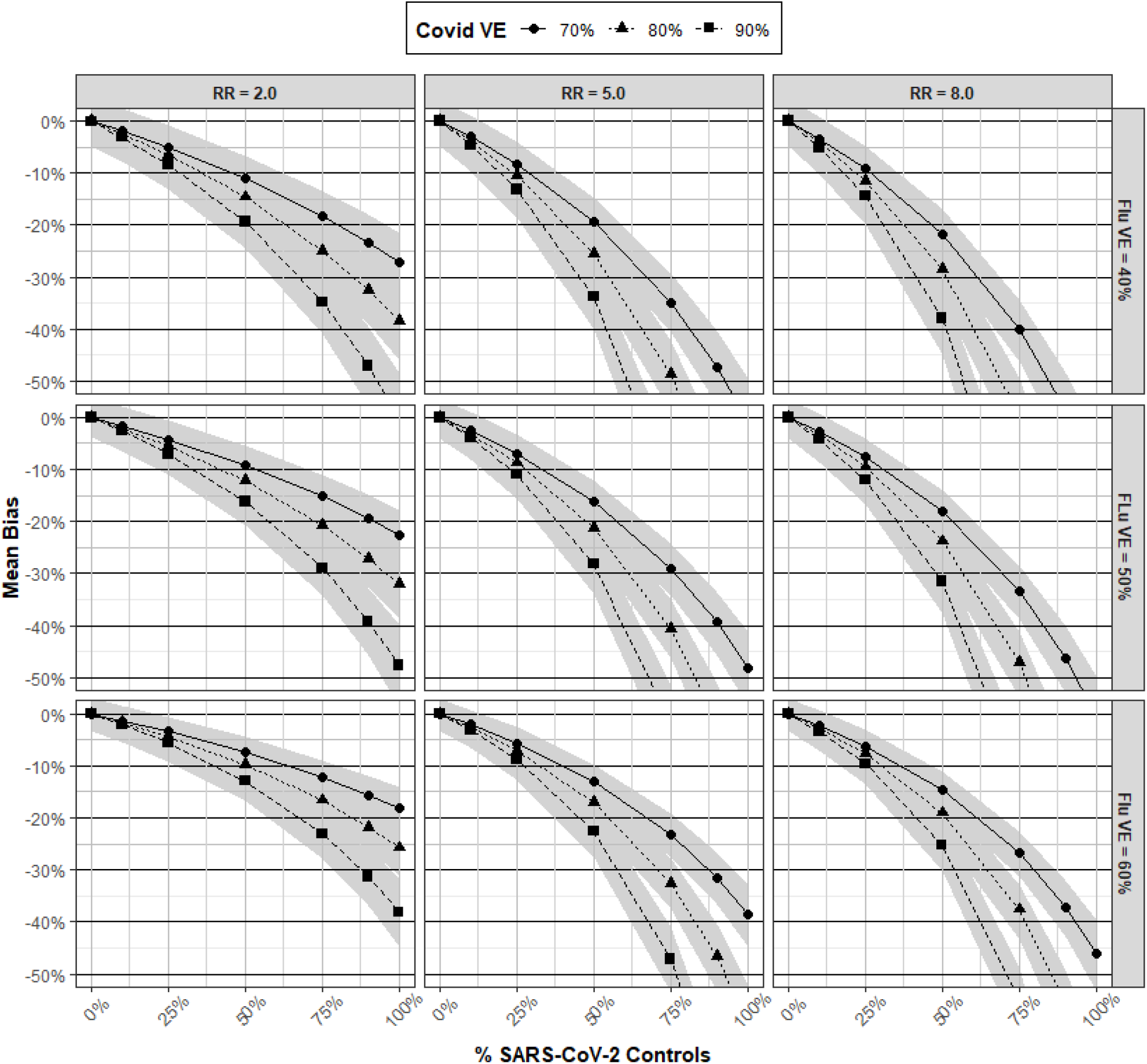
Mean bias and 95% confidence intervals (CI) in influenza vaccine effectiveness (VE) estimates derived from a test-negative study with SARS-CoV-2 controls under varying scenarios of *RR*_*flu vx*|*covid vx*_, *VE*_*flu*_, *VE*_*covid*_, and *P*_*controls*_.

**Table 1.** Input parameter values for simulations. Where multiple values are specified, populations were simulated for each value separately.

| Parameter | Description | Values |
| --- | --- | --- |
| $Cov_{flu}$ | Flu vaccination coverage | 55% |
| $Cov_{covid}$ | COVID-19 vaccination coverage | 70% |
| $IP_{flu}$ | Incidence proportion of medically-attended influenza ARI among persons unvaccinated for influenza | 5% |
| $IP_{covid}$ | Incidence proportion of medically-attended COVID-19 ARI among persons unvaccinated for COVID-19 | 5% |
| $VE_{flu}$ | Influenza vaccine effectiveness | 40%, 50%, 60% |
| $VE_{covid}$ | COVID-19 vaccine effectiveness | 70%, 80%, 90% |
| $P_{controls}$ | Proportion of controls represented by vaccine-preventable disease of interest | 0, 0.1, 0.25, 0.5, 0.75, 0.9, 1.0 |
| $RR_{covid\ vx\vert flu\ vx}$ | Risk ratio comparing the uptake of COVID-19 vaccination among persons receiving and not receiving influenza vaccination | 1.5, 2.0, 3.0 |
| $RR_{flu\ vx\vert covid\ vx}$ | Risk ratio comparing the uptake of influenza vaccination among persons receiving and not receiving COVID-19 vaccination | 2.0, 5.0, 8.0 |

### Bias in influenza VE estimates

In all scenarios, the inclusion of SARS-CoV-2 controls underestimated influenza VE. In general, bias in influenza VE estimates was higher than in COVID-19 studies. However, patterns of bias in influenza VE estimates were similar to those in COVID-19 studies, where greater bias was observed with lower values of *VE*_*flu*_ (*i*.*e*., the vaccine of interest), higher *VE*_*covid*_ (*i*.*e*., the confounding vaccination), increasing *RR*_*flu vx*|*covid vx*_, and increasing *P*_*controls*_. While bias was low for all scenarios that included ≤10% of SARS-CoV-2 *P*_*controls*_, there was moderate bias in some scenarios where SARS-CoV-2 *P*_*controls*_ approached 25%. High bias was observed for several scenarios with 50% *P*_*controls*_ of SARS-CoV-2, and all scenarios had moderate to high bias with ≥75% *P*_*controls*_ of SARS-CoV-2.

## Discussion

In this paper, we provide the theoretical basis and quantification of confounding bias in COVID-19 and influenza VE test-negative designs related to the inclusion of influenza and SARS-CoV-2 controls, respectively. While positive correlation in the uptake of influenza and COVID-19 vaccination consistently led to the underestimation of VE, there was minimal bias in scenarios that included low percentages of controls with the other vaccine-preventable ARI. Specifically, bias in COVID-19 VE estimates was low for all scenarios with ≤50% of influenza controls. For influenza VE test-negative designs, bias was low in all scenarios with ≤10% of SARS-CoV-2 controls. Where the proportions of controls with the other vaccine-preventable ARI exceeds these levels, moderate to high bias in VE estimates can occur.

Although confounding in influenza and COVID-19 VE estimates is driven by correlated vaccine behaviors, we found the magnitude of bias was highly dependent upon the true VE of both the vaccine of interest and the confounding vaccination. In general, where the true VE was high for the vaccine of interest and low for the confounding vaccine, there was less bias in VE estimates. Since true influenza VE is expected to be lower than true COVID-19 VE,^4, 18-22^ this relationship exacerbates bias in influenza VE studies, which are already more likely to have higher bias based on a greater conditional probability of vaccination. Particularly in the setting of the COVID-19 pandemic, where influenza VE test-negative controls may be more likely to represent persons with SARS-CoV-2, these relationships suggest researchers should consider deconfounding methods to avoid meaningful bias in influenza test-negative VE estimates.

While our findings may be viewed as reassuring regarding bias in COVID-19 test-negative VE studies, these results are subject to several important limitations. First, we caution that our simulations examine bias scenarios we considered likely in the general population based on empiric U.S. data; however, we did not explore plausible scenarios by subpopulation. It is possible that some subpopulations, such as older persons or persons who are at higher risk of severe disease, may have a higher conditional probability of COVID-19 vaccination, given influenza vaccination status; this difference could thus, cause greater bias than we observed in our estimates. Similarly, regional variation in COVID-19 and influenza vaccination coverage may also affect the conditional probability of vaccination. Currently, lower uptake of COVID-19 vaccination in some southern U.S. states more closely aligns with influenza vaccination coverage in these regions.^16, 17^ These differences may also increase the conditional probability of vaccination, and thus, represent a setting where greater bias in COVID-19 VE estimates can occur. Both examples demonstrate that bias in VE estimates may be differential by subpopulation, which may be important for the interpretation of VE results. Additionally, even where the conditional probability of vaccination is the same, we found that bias can vary by true VE. In the case of COVID-19, where true VE is likely to vary by vaccine product,^4, 20^ bias in VE estimates will be differential by vaccine product. For similar reasons, bias in COVID-19 VE estimates may also be differential by outcome, such as symptomatic disease versus hospitalization. Collectively, these examples may highlight the importance of adopting deconfounding methods to promote comparability in VE estimates across subpopulations or outcomes, even in situations where overall bias is expected to be low.

It is important to acknowledge a deconfounding requirement may impact the feasibility of a test-negative study, *i*.*e*., a design commonly implemented using administrative data.^2, 4^ In particular, a challenge of deconfounding in the analysis is that it requires measurement of both vaccinations, which may not both be reliably recorded in a vaccine registry. Further, deconfounding by study design requires the additional costs associated with testing for other pathogen(s), unless this testing is routinely performed. However, where additional efforts are made to implement these methods, an advantage is that both influenza and COVID-19 test-negative VE studies can be run in parallel without much extra additional effort.

In conclusion, our work suggests bias is low in VE estimates derived from COVID-19 and influenza test-negative studies that include influenza and SARS-CoV-2 controls, respectively, in situations where these controls represent a low proportion of total test-negative controls. Nonetheless, we encourage researchers to thoughtfully consider this potential bias and its implications in their respective study settings. Where researchers determine bias is unlikely to be meaningful and do not undertake deconfounding methods to remove bias, adequate justification should be provided to promote critical interpretation and confidence in study results.

## Data Availability

The datasets that were used to inform the marginal probabilities of influenza and COVID-19 vaccination were derived from sources in the public domain.

## Author contributions

MKD designed the study and drafted the article. SMP helped to interpret the findings and critically reviewed the manuscript. AV and JM directed the analytical strategy, helped to interpret the findings, and critically reviewed the manuscript.

## Data availability

The datasets that were used to inform the marginal probabilities of influenza and COVID-19 vaccination were derived from sources in the public domain.^14^

## Supplementary data

Supplementary data are available in Supplemental Appendix 1.

## Supplemental Appendix 1

### Estimation of the conditional probabilities of COVID-19 and influenza vaccination

Since bias in VE estimates is driven by the relationship between COVID-19 and influenza vaccine uptake, we examined the conditional probabilities of COVID-19 and influenza vaccination using data from U.S. IPSOS Knowledge Panel and NORC AmeriSpeak Omnibus Surveys.^14^ Briefly, the U.S. Centers for Disease Control and Prevention sponsored questions on these surveys pertaining to influenza and COVID-19 vaccination coverage and intent.^14^ Both surveys aimed to recruit nationally representative panels of the U.S. population, and are administered twice per month to assess national influenza vaccination coverage and intent among U.S. adults.^14^ We used publicly-available data from these surveys collected from September 10-13, 2021 for our analyses, which represented the most recent survey data at the time of our analysis. Survey questions asked participants about their COVID-19 vaccination uptake and intent, as well as their influenza vaccination uptake and intent. Specifically, for COVID-19 vaccination, three categories of uptake/intent were provided: (i) vaccinated or definitely plan to get vaccinated for COVID-19, (ii) probably will get vaccinated for COVID-19 or are unsure, and (iii) probably or definitely will not get vaccinated for COVID-19. For influenza vaccination, four categories of uptake/intent were provided: (i) vaccinated for flu, (ii) intend to get vaccinated for flu, (iii) not sure about getting vaccinated for flu, and (iv) do not intend to get vaccinated for flu.

To understand the impact of correlated COVID-19 and influenza vaccination behaviors, we estimated the conditional probabilities of: (i) COVID-19 vaccination, given influenza vaccination status, and (ii) influenza vaccination, given COVID-19 vaccination status using survey data. We characterized each of these conditional probabilities as a risk ratio, where:

i. *RR*_*covid vx* | *flu vx*_ = risk ratio comparing the uptake (risk) of COVID-19 vaccine between persons who did and did not receive influenza vaccination
ii. *RR*_*flu vx* | *covid vx*_ = risk ratio comparing the uptake (risk) of flu vaccine between persons who did and did not receive a COVID-19 vaccination

Given that answer categories for influenza or COVID-19 vaccination survey questions did not represent participant’s definitive influenza and COVID-19 vaccination status, we estimated *RR*_*covid vx*|*flu vx*_ and *RR*_*flu vx*|*covid vx*_ using two different definitions for COVID-19 vaccine uptake and two different definitions for influenza vaccine (Supplemental Table 1). For each combination of COVID-19 and influenza definitions (total n = 4), we used survey point estimates to estimate the marginal probabilities of influenza and COVID-19 vaccination. We then applied these probabilities to a population of approximately 200,000, to estimate the marginal vaccination status for the population. For each scenario, *RR*_*covid vx*|*flu vx*_ and *RR*_*flu vx*|*covid vx*_ were estimated. A range of 1.5 to 1.9 was estimated for *RR*_*covid vx*|*flu vx*_; a range of 2.4 to 8.3 was estimated for *RR*_*flu vx*|*covid vx*_. Based on these results, we selected *RR*_*covid vx*|*flu vx*_ values of 1.5 and 2.0, and *RR*_*flu vx*|*covid vx*_ values of 2.0, 5.0, 8.0 for our simulation inputs.

**Supplemental Table 1.** Definitions for influenza vaccination and COVID-19 vaccination applied to survey data.

| **Disease** | **Definition #** | **Survey categories**<br>**used to define vaccination status = Yes** |
| --- | --- | --- |
| **Influenza** | 1 | a) vaccinated for flu, *OR*<br>b) intend to get vaccinated for flu |
|  | 2 | a) vaccinated for flu, *OR*<br>b) intend to get vaccinated for flu, *OR*<br>c) not sure about getting vaccinated for flu |
| **COVID-19** | 1 | a) vaccinated or definitely plan to get vaccinated for COVID-19 |
|  | 2 | a) vaccinated or definitely plan to get vaccinated for COVID-19, *OR*<br>b) probably will get vaccinated for COVID-19 or are unsure |

